# Basic estimation-prediction techniques for Covid-19, and a prediction for Stockholm

**DOI:** 10.1101/2020.04.15.20066050

**Authors:** Tom Britton

## Abstract

Predicting future infections for covid-19 is essential in planning healthcare system as well as deciding on relaxed or strengthened preventive measures. Here a quick and simple estimation-prediction method for an urban area is presented, a method which only uses the observed initial doubling time and *R*_0_, and prediction is performed without or with preventive measures put in place. The method is applied to the urban area of Stockholm, and predictions indicate that the peak of infections happened in mid-April and infections start settling towards end of May.

## Introduction

The covid-19 is currently spreading at rapid pace in most countries of Europe, resulting in high number of case fatalities and healthcare systems being overwhelmed. We here present a quick and simple method for predicting the progress of the epidemic in a community which is fairly well-mixed, an urban region being a typical example. The simplicity of the method makes model assumptions more transparent and it is easier to study which parameters are most influential to the predictions and studying how they affect predictions. The simple method should preferably be complemented with more complex and realistic modelling, perhaps on a national scale [1,2].

## Method summary

The methodology is described in detail in the supplementary material; here is a very short description. The initial doubling time *d* and the basic reproduction number *R*_0_ are used to parametrize the General epidemic model [3]. Calibration to calendar time is done using the observed number of case fatalities, together with estimates of the time between infection to death, and the infection fatality risk. Finally, predictions are made assuming no change of behaviour, as well as for the situation where preventive measures are put in place at one specific time-point. The overall effect of the preventive measures is assumed to be known, or else estimated from the observed increased doubling time after preventive measures are put in place.

## Illustration: Predicting the outbreak in Stockholm

We illustrate our methods on the Stockholm region in Sweden. Greater Stockholm urban area has around *N*=2 million people, and the initial doubling time of cumulative case fatalities was around *d*=3.5 days before preventive measures were put in place [4], and we assume that R_0_=2.5 this being a common value [5]. A number of (mainly but not exclusively voluntary) preventive measures were put in place around the date *t_p_=* March 16. We assume the typical time between infection and death equals *s_D_*=21 days. These preventive measures will start affecting fatality rates at *t_p_* + *s_D_* = April 6. We are now (=April 26) 2.5 weeks later so the effect is uncertain, but a new longer effective doubling time of *d_E_*=14 days seems to be a reasonable estimate based on case fatalities between April 9 and April 23 where the total number has close to doubled [4]. To calibrate the relative time to calendar time we use that the cumulative number of case fatalities on March 31, before effect of preventive measures, equals 150. We emphasize that the quantities are by no means precisely estimated, so results contain a lot of uncertainty and are mainly an illustration of the method.

These quantities are used to generate an epidemic and also to calibrate it to calendar time, as described in the supplementary material. Once the calibration is performed it is possible to predict future infections in the outbreak, both with and without preventive measures of different magnitudes. The results are summarized in Figure 1 and Figure 2 below. Figure 1 reports the daily incidence of new infections for the different scenarios.

**Figure 1:**
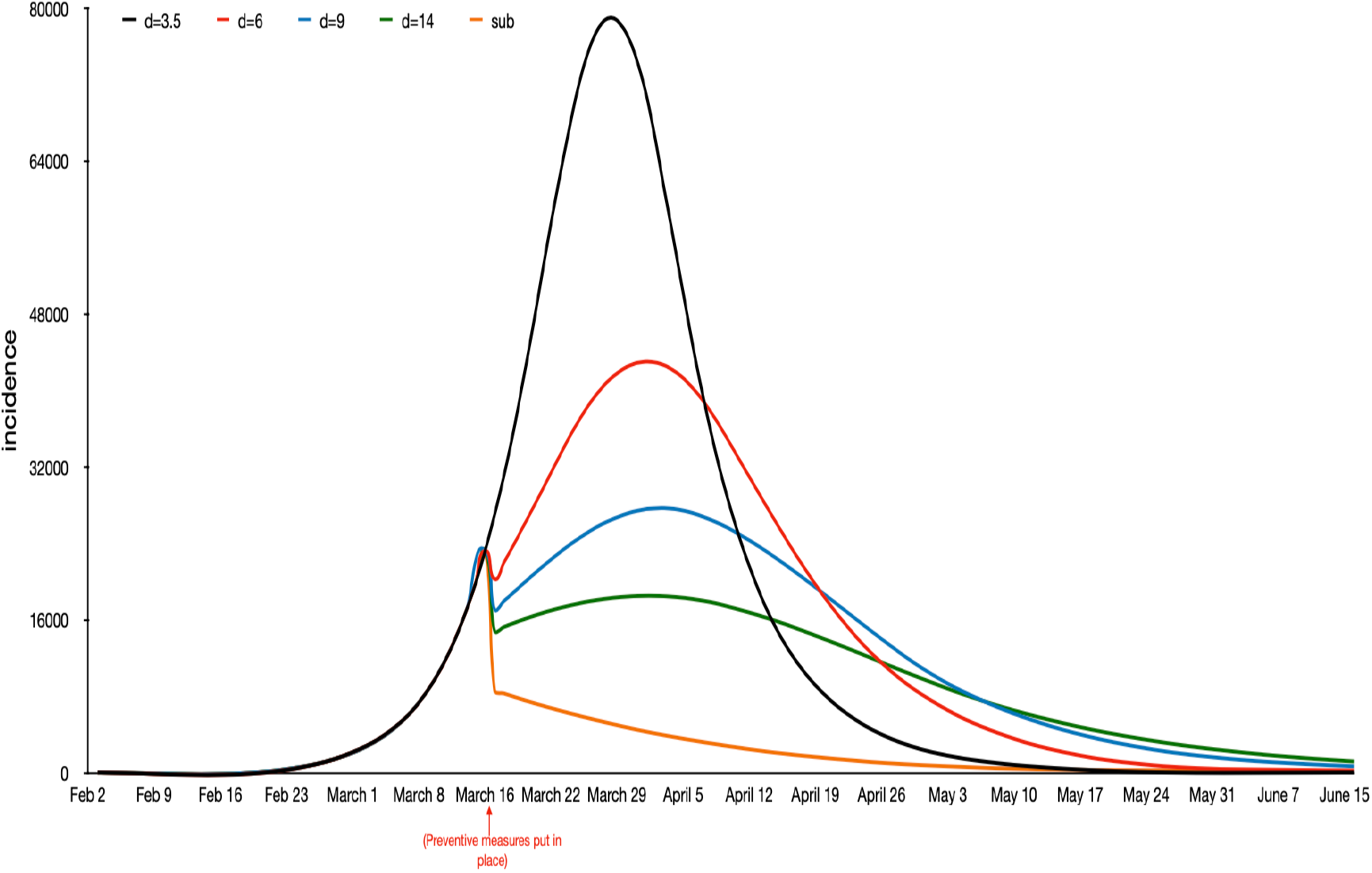
Plot of the daily number of new infections over time, with preventive measures, and for a variety of magnitude of preventive measures.

**Figure 2:**
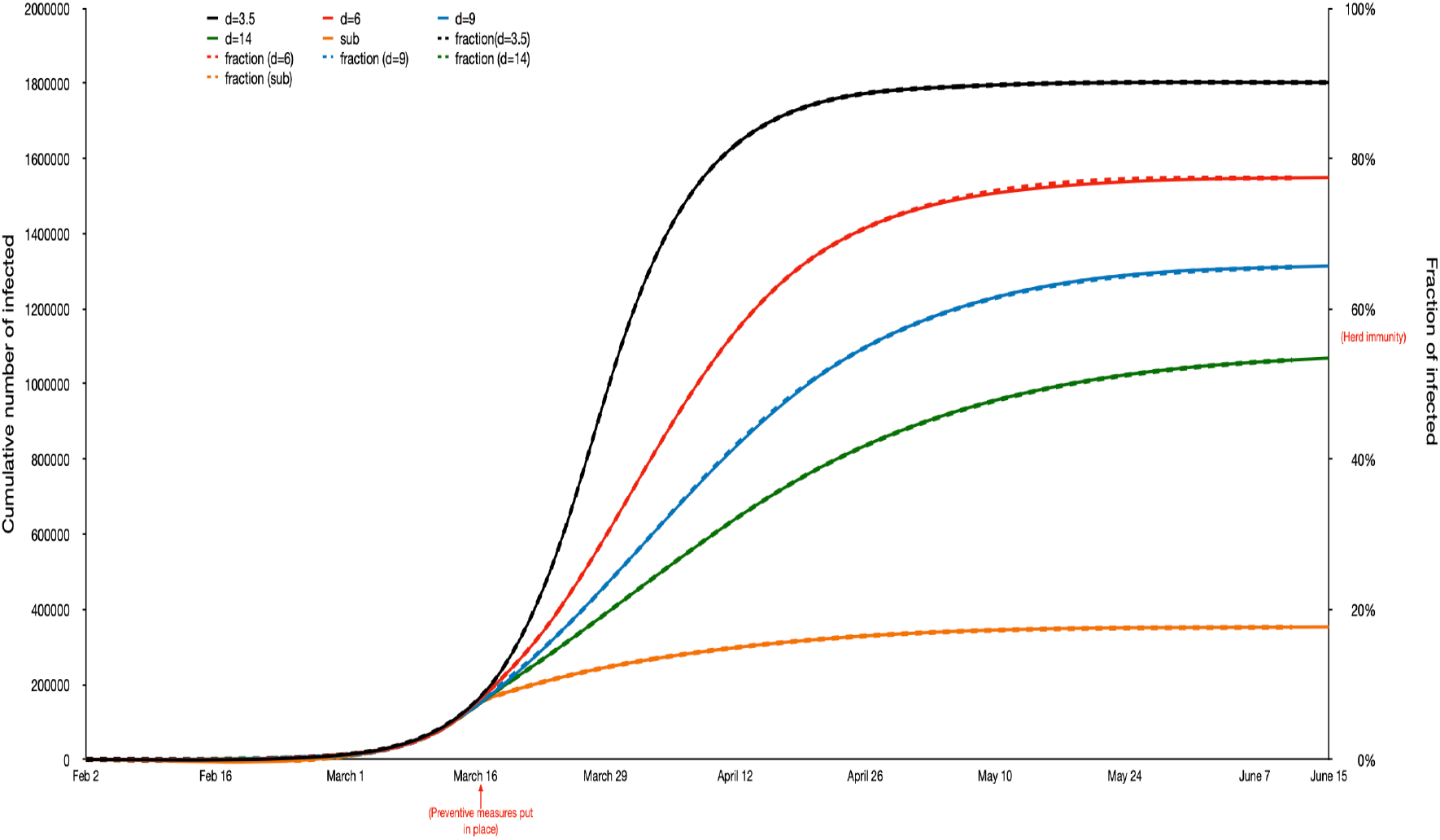
Plot of the cumulative number infected over time. Note that the two preventive measures with largest magnitude do not reach herd immunity.

**Caption:** Plot of the predicted daily number of new infections over time in Stockholm region, without and with preventive measures of different magnitude.

As described in the Supplementary material a better approximation, taking heterogeneities into account, is to flatten the curve by 25% and to shift it to the right 1.5 weeks (the left part 1 week and the right part 2 weeks). If this is taken into account, and we assume the new doubling time equals *d_E_*=14 days (green curve), suggests that the peak of transmission happened around April 11, 75% of all infections will have happened by May 7 and 90% of all infections will have happened May 28 (assuming preventive measures remain). Needless to say, there is of course uncertainty in these time points – the true time points may occur one or even two weeks earlier or later. Comparing the different scenarios we see that the dates do not change dramatically for different scenarios except for the yellow curve corresponding to severe lock-down. The main difference between the other curves is the overall height, and that the right tail is heavier the more effective preventive measures are. Had the preventive measures happened earlier in relation to the outbreak, as for example was the case with other parts of Sweden, the peak heights would have been lower and shifted further forward in time.

Figure 2 shows the cumulative numbers of infected (with overall percentages to the right) for the different scenarios.

**Caption:** Plot of the predicted cumulative number infected over time in Stockholm region, both without and with preventive measures of different magnitude. Note that the two preventive measures with largest magnitude do not reach close to herd immunity.

As described above the increase in the curve should be shifted to the right to fit better reality. As for the end of the curves they should be lowered by 15%. Without preventive measures, taking this correction into account, we see that approximately 77% would get infected. And for our main prediction, the green curve predicts that 47% will get infected (assuming preventive measures remain). An important observation is that in the first two scenarios, the final fraction infected exceeds the critical immunity level ν__C_=1-1/ *R_0_*=60% [3]. This implies that in these two situations the community has reached herd immunity and is hence protected from additional outbreaks when preventive measures are relaxed (assuming infection induce complete immunity!). The blue curve (when deflated by 15%) is just below herd immunity, and the two latter scenarios, our main prediction with new doubling time of *d_E_=14* days, and the situation where preventive measures give *R_E_*=0.80, the final fraction infected is clearly below herd immunity. Fewer infected is of course a big gain. However, since the corresponding fractions lie below herd immunity, the community is at risk for additional outbreaks if all preventive measures are relaxed. This is particularly the case for the last scenario where only 15% get infected during the outbreak. We emphasize that our method is an approximation equipped with several uncertainties which are discussed more in the Supplementary material.

The Swedish Public Health Agency recently predicted infections of covid-19 in the Stockholm region [6]. There it was predicted that the fraction infected by May 1 will be 26\%. The green curve in our analysis gives a very similar prediction: shifting the curve 2 weeks forward corresponds to observing the green curve April 17, and after reducing by 15%, the green curve predicts 27% infected. Our prediction may also be compared with earlier published estimates for Sweden [7] where it was predicted that by March 28, 3.1% of the Swedish population would be infected (with credibility bound 0.85%-8.4%). Our best prediction (as described above) would be to look one week earlier, so March 21, and the green curve of Figure 2 multiplied but 0.85, which gives 10.5% infected. We note that our prediction is for Stockholm which had the vast majority of all infections in the beginning of the outbreak. And, since the Stockholm region makes up 20% of the country population, 10.5% infected in Stockholm could very well agree with 3.1% (or slightly more) in all of Sweden.

## Conclusions and Discussion

We have demonstrated a quick and simple method to estimate and predict an on-going epidemic outbreak both with and without preventive measures put in place. As input data we use the basic reproduction number *R*_0_ and the doubling time during the early stage of the epidemic, and its new doubling time after preventive measures are put in place. The method also uses the reported cumulative number of deaths at a given time, the typical time between infection and death *s_D_*, and the infection fatality risk *f*, in order to time-calibrate the model to calendar time. The method is most sensitive to the doubling times, to some extent also to *R*_0_, but less sensitive to the latter quantities which are often equipped with high uncertainty. The main purpose of the present paper is to use it as a complement to more advanced models on a national scale, models where it is often less clear which parameters have biggest influence on the conclusions.

In the current paper the focus has been on predicting the number of infected over time. Clearly, the burden on the healthcare system, measured by hospitalized patients or case fatalities, is more important. When predicting these quantities, an age-structured model is advantageous since the risk for severe symptoms and death increases with age. There is however high uncertainty in what fraction of all infected that will require healthcare and that will die for different age-groups, as well as the overall infection fatality risk *f*. For example, current estimates of *f* (not to be mixed up with case fatality risk, cfr) vary between 0.2% up to 1% (e.g. [8], [9]). Clearly, any prediction of the number of fatalities will be equipped with very large uncertainty due to uncertainty in *f*, let alone all other uncertainties, and this big uncertainty is most often not acknowledged by instead picking just one published estimate of *f*.

## Data Availability

I have only used publicly available data

## Supplementary material: Basic estimation-prediction techniques for Covid-19, and a prediction for Stockholm

### 1 Introduction

We here give a) the details for the estimation-prediction method mentioned in the main paper, b) some more details on the illustration prediction of Stockholm, and c) some additional discussion points. The method is applicable for a community which is fairly well-mixed, an urban region being a typical example.

We focus on predicting of the main phase of the epidemic, and not on the very beginning or end (when very few are infectious implying that randomness plays a crucial role). For this reason we use a deterministic epidemic model. More specifically we use the General epidemic model [4] because it allows for more heterogeneity in how many infectious contacts different infected have, and when in time these happen, in comparison to generation-time models such as the Reed-Frost model [4]. The model hence allows for heterogeneity in terms of infections, but all individuals are equally susceptible and mix homogeneously.

As input to our prediction model we use the observed doubling time *d* during the initial (random) phase of the epidemic and the basic reproduction number *R*_0_. Estimation of *d* is straightforward (e.g. [1]) and many estimates of *R*_0_ for covid-19 can be found in the literature, most of them lying in the range 2.2-2.8 (e.g. [9], [8]). We start by describing the different steps in the methodology, including also how to time-calibrate the model to calendar time, and then apply our method by predicting the Covid-19 outbreak in the Stockholm region.

### 2 Methods

We now present our estimation-prediction procedure. We start by predicting the behaviour of the outbreak based on knowing the basic reproduction number *R*_0_ and the doubling time *d* in the initial phase of the epidemic. We then time-calibrate the model in the sense of estimating where in the epidemic outbreak we are on a given date *t*_1_ by using cumulative case fatalities and knowledge of the typical time between infection and death *s_D_*. Finally, we predict the time calibrated epidemic outbreak under the assumption that a set of preventive measures are put in place a given date *t_p_* during the early phase of the epidemic outbreak. This is done either assuming the overall reduction in spreading is known, or else by assuming that the increased doubling time after prevention, *d_p_*, is observed.

#### 2.1 Prediction based on *R*_0_ and initial doubling time *d*

As input for our prediction we use the observed doubling time *d* during the initial growth rate and the basic reproduction number *R*_0_, both valid before any preventive measures were put in place. The doubling time can for example be estimated from the empirical doubling time of case fatalities as described in [1]. The doubling time relates to the exponential growth rate *r* of the epidemic by the relation *e^rd^* = 2, so *r* = ln(2)/*d*. The basic reproduction number can be estimated using other data sources, for example using contact tracing giving information about the (random) generation time *G* and its mean *g* = *E*(*G*) (this is however not trivial, cf. [2], [12]). Here we assume the initial doubling time d and the basic reproduction number *R*_0_ to be known.

We assume the epidemic progresses according to the General epidemic model (GEM) [4]. For this model, the mean generation time relates to *r* and *R*_0_ by the relation [11]

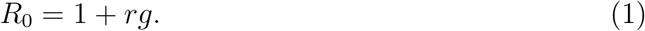

We hence have that the mean generation time equals *g* = (*R*_0_ – 1)/r. The GEM has two model parameters: the rate of infectious contact A that infectious individuals have, and the rate of recovery *γ* (e.g. [4]). The basic reproduction number for GEM equals *λ*/*γ* (the rate of infectious contacts multiplied by the mean duration of the infectious period 1/*γ*. The rate of infecting someone s time units after infection is *λe*^−^*^γs^*, and since the mean number of contacts equals *R*_0_ = *λ*/*γ*, it follows that *G* has density *f_G_*(*s*) = *λe*^−^*^γs^*/(*λ*/*γ*) = *γe*^−^*^γs^*. It hence follows that *G* is exponential with parameter *γ* with mean *E*(*G*) = 1/*γ*.

From our input data, *d* and *R*_0_, we hence conclude that *g* = (*R*_0_ − 1)*d*/ ln(2) and hence that *γ* = ln(2)/(*d*(*R*_0_ − 1)).

The contact rate *λ* can also be obtained from our input values *R*_0_ and *d* from the fact that *R*_0_ = *λ*/*γ* for the GEM [4]. We immediately have that *λ* = *R*_0_*γ* = (*R*_0_/(*R*_0_ − 1))(ln(2)/*d*).

Once we have calibrated our parameters *λ* and *γ* to *R*_0_ and the observed initial doubling time *d* we simply use the GEM to predict the epidemic (assuming no preventive measure are put in place). Suppose the community size is *N* (assumed large) and that we start with a small fraction (but fairly large number) infected and the rest being susceptible, for example *i*_0_ = 50 *s*_0_ = *N* − 50, where *s_t_* and *i_t_* denote the number of susceptible and the number of infectious individuals respectively (the index *t* is hence relative to the start of the epidemic and not calendar time). The transitions of the GEM are given by

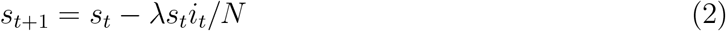

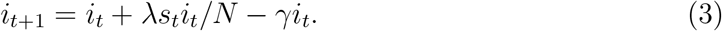

The interpretation is that each infectious individual at time *t* has on average *λ* infectious contacts per day, and with probability *s_t_*/*N* each such contact results infection. Those susceptibles who get infected move to the infectious state. The other transition is for an infectious individual to stop being infectious and recovering and becoming immune (a few also die). The number of individuals who have recovered (also including the few who die) equals *N* − *s_t_* − *i_t_*. This system can be iterated forward sequentially until the first time *T* when the number of infectious individuals drops below 1: *i_T_* ≤ 1 (it never reaches exactly 0 but asymptotes to 0). The number of individuals who have been infected by *t* equals *N* − *s_t_*.

By solving the iterative system it is easy to plot the number of infectives *i_t_* and the number of infected *N* − *s_t_* over time, as well as the daily incidence *s_t_*_−1_ − *s_t_*.

#### 2.2 Time calibration based on initial case fatality data

In the previous subsection an epidemic model was fitted to the observed initial doubling time *d* and the known basic reproduction number *R*_0_. An important remaining task is to identify where in this outbreak the epidemic is at calendar time *t*_1_ (’’today”) (cf. [1]). We hence want to know which relative time *t* since the start of the epidemic that corresponds to the calendar time *t*_1_. In order to do so we use the observed total number of case fatalities up to calendar time *t*_1_, denoted Λ(*t*_1_). We further need approximate knowledge of two quantities: the typical duration *s_D_* between getting infected and dying (for those who die) and the infection fatality ratio f being defined as the probality that an individual who gets infected dies. For covid-19 *s_D_* ≈ 21 days but estimates of *f* vary in the range 0.2% − 1% (e.g. [13], [12]). Fortunately it turns out that the time calibration is not too sensitive to these numbers.

Given Λ(*t*_1_), *s_D_* and *f* we know that the Λ(*t*_1_) who have died by *t*_1_ were all infected by *t*_1_ − *s*_D_. But these ”to-die” infected individuals only make up a fraction *f* of all who were infected by *t*_1_ − *s*_D_, so the number of infected individuals at calendar time *t*_1_ − *s_D_* equals Λ(*t*_1_)/*f*. This means that we calibrate calendar time to time relative to the start of the epidemic by equating *t*_1_ — *s_D_* to the relative value *t*^*^ for which 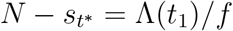.

Given this time calibration between relative and calendar time, *t*^*^ = *t*_1_ − *s_D_*, the estimated number of infected people at present time equals 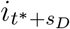 and e.g. the predicted number of infectious individuals three weeks later (calendar time *t*_1_ + 21) equals 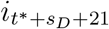.

#### 2.3 Prediction with preventive measures put in place

Suppose that a set of preventive measures are put in place at some calendar time *t_p_* still assumed to be in the early phase of the epidemic. Here we assume that this affects the rate of infectious contacts but not the (mean) generation time *g* = 1/*γ*. Most preventive measures agree with this: school closure, self-isolation, closing (or reduced activities) of restaurants, bars, cinemas. There are also some preventions which aim at reducing *g*, such as contact tracing followed by isolation, but here we restrict ourselves to preventions reducing λ. We assume that the new preventive measures have the overall effect of reducing λ by a factor *ρ*, so that the new *effective* rate of contact equals λ*_E_* = λ(1 − *ρ*), and the new effective reproduction number equals *R_E_* = (1 − *ρ*)*R*_0_. For covid-19 there is currently no available vaccine, but for situations where there is it is also possible to include vaccination of a fraction of the community as a preventive measure. In this case, the factor *ρ* also includes effects from vaccination. If for example a fraction *ν* are vaccinated with a vaccine giving perfect immunity then this results in *ρ* = *ν* if this is the only preventive measure.

In applications it is close to impossible to know the overall magnitude of the preventive measures *ρ* when a set of preventive measure are put in place jointly. In [1] it is shown how to estimate *ρ* by observing the change in the doubling time once the preventive measures have influenced e.g. fatality rates. Applying Equation (3) in [1] it follows that

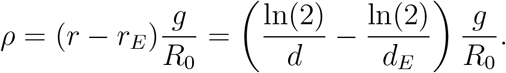

Above *r* is the exponential growth rate before preventive measures and *r_E_* is the growth rate after preventive measures have affected case fatality rates, and similarly for the doubling times *d* and *d_E_*.

The preventive measure hence induce a lower growth rate *r_E_* = ln(2)/*d_E_*. We assume the same mean generation time *g* = 1/*γ*, and since *r_E_* = *λ_E_* − *γ* it follows that the new contact rate *λ_E_* = *r_E_* + *γ*. The new reproduction number equals *R_E_* = 1 + *r_E_g* = 1 + *r_E_*/*γ*.

To sum up, if *R*_0_ and *r* = ln(2)/*d* are known from before preventive measures, and preventive measures are put in place on day *t_p_* resulting in a new longer doubling time *d_E_*, then the time calibrated prediction model should change from *λ* = (ln(2)/*d*)*R*_0_/(*R*_0_ − 1) to *λe* = (ln(2)/*d_E_*) + *γ*.

#### 2.4 An improved approximation

The method is developed for an urban area because the underlying methodology assumes a homogeneous community. Even for an urban area there are many heterogeneities, both in terms of susceptibility and infectivity, but in particular with respect to how people interact. The effect of such heterogeneities is that the outbreak is slower, the peak is delayed and lower, and fewer people get infected. The magnitude of these effects depend on the particular community, and a thourough investigation would require a much more advanced model, which the current method wants to avoid. For this reason we suggest to use the following corrections to make our current simple model fit better to real life epidemics in an urban area.

##### Modification

Shift the curve of new infections (corresponding to Fig 1) 1-2 weeks forward in time: the left part of the curve can be shifted 1 week, the peak 1.5 weeks and the right part 2 weeks. Reduce the size of the peak by 25%, and finally, reduce the fraction getting infected at the end of the outbreak (Fig 2) by 15%.

### 3 Predicting the outbreak in Stockholm

We illustrate our methods on the Stockholm region in Sweden. Greater Stockholm urban area has around *N* = 2 million people, and the initial doubling time of cumulative case fatalities was around *d* = 3.5 days before preventive measures were put in place [7], and we assume that *R*_0_ = 2.5 this being a common estimate [9]. A number of (mainly but not exclusively voluntary) preventive measures were put in place around the date *t_p_* = March 16. We assume the typical time between infection and death (for those who die from covid-19) equals *s_D_* = 21 days. These preventive measures will start affecting fatality rates around *t_p_* + *s_D_* = April 6. This analysis is performed April 26, with reliable fatality data up to about April 23. The number of case fatalities have just about doubled between April 9 and April 23 [7], so we estimate the effective doubling time to *d_E_* = 14. To calibrate the relative time to calendar time we finally assume that the cumulative number of case fatalities on March 31, before effects of preventive measures, equals Λ(March 31) = 150. We emphasize that the quantities are by no means precisely estimated, so results contain a lot of uncertainty.

To start, the evolution of the epidemic outbreak is simulated using the system defined above with *λ* = (ln(2)/3.5)*2.5/1.5 = 0.330. The rate of recovery equals *γ* = *λ* − ln(2)/*d* = 0.132. Finally, the new contact rate *λ_E_* induced by the new bigger doubling time *d_E_* = 14 equals *λ_E_* = (ln(2)/*d_E_*) + 7 = 0.182. As a consequence, the iterative prediction model should have *λ* = 0.330 replaced by *λ_E_* = 0.182 on March 16 and onwards. As a side remark we note that this change of doubling time from 3.5 days to 14 days corresponds to changing *R*_0_ = 2.5 to *R_E_* = 1 + (ln(2)/*d_E_*)/*γ* = 1.27 giving the magnitude of preventive effects of *ρ* = 1 − *R_E_*/*R*_0_ = 0.49 so close to 50% overall reduction in contact rates. Since the estimate *d_E_* = 14 days of the new doubling time (after preventive measures) is uncertain we do similar calculations had the new doubling time instead been 6 and 9 days respectively, and also for the situation where the preventive measures reduces *R_E_* to 0.8 implying that it directly starts decaying.

Finally the time calibration. For this we set the infection fatality risk to *f* = 0.3% as a guess. As mentioned above it will not change the time calibration more than a week if the true fatality risk is 0.1% or 1%. The 150 case fatalities by March 31 would hence imply that the number of infected three weeks earlier, March 10, equals 200/0.003 = 50 000. We therefore calibrate March 10 to the relative day t at which the cumulative number of infected equals 50 000 (or as close to as possible). This turns out to be on day *t* = 31 of the epidemic. The estimate that 50 000 were infected in Stockholm by March 10 might seem high, since most of the transmission is believed to have been imported in late February when many families returned from skiing in the Italian alps, but the number infected less than two weeks earlier should not have exceeded 1000 people or so. However, there were many flights to Stockholm from Milan as well as China during all of february, most likely bringing transmission into Sweden also earlier, so it is not unreasonable that the number of infected by March 10 could be at least close to 50 000.

Once the calibration is performed it is possible to predict relevant quantities of the outbreak, both with and without preventive measures of different magnitudes. The results are summarized in Figure 1 and Figure 2 below. Figure 1 reports the daily incidence of new infections for the different scenarios. Recall the suggested approximation improvements of Section 2.4. For example, had no preventive measures taken place (black curve) the peak day when most infections would have taken place is April 7 (March 28 + 10 days), and this day around 59 000 people would have been infected (79 000 multiplied by 0.75). For our main prediction (green curve) the peak day of transmission is April 11 (April 1 + 10 days). The first peak March 15, just before preventive measures were put in place, is partly an artefact from the model assuming all preventive measures were initiated on one single day. The height of the green peak is dramatically lower than the black curve. Instead of 59 000 people infected on the peak day the green curve has 14 000 infected.

Comparing the different scenarios, without preventions and with preventions of different magnitude, it is seen that the peak is reduced the higher magnitude of preventive measures, and slightly shifted forward in time. Had the preventive measures happened earlier in relation to the outbreak, as for example was the case with other parts of Sweden, the peak heights would have been lower and further shifted forward in time.

Figure 2 shows the cumulative numbers of infected (with overall percentages to the right) for the different scenarios. Using the improved approximation it can be seen that for the main prediction (green curve) 75% of all infections have taken place by May 7 and 90% of all infections will have ocurred by May 28 (assuming preventive measures remain).

Another important observation is that in the first two scenarios, the final fraction infected exceeds the critical immunity level *ν*_C_ = 1 − 1/*R*_0_ = 0.6 [4], and the blue curve (multiplied by 0.85) reaches very close to herd immunity. This implies that in these three situations the community has reached herd immunity and is hence protected from additional outbreaks when preventive measures are relaxed (assuming infection induce complete immunity!). For the latter two scenarios, our main prediction with a new doubling time of *d_E_* = 14 days, and the situation where preventive measures give *R_E_* = 0.80, the final fraction infected are both below 50% which of course is positive compared to the upper three curves. However, since the corresponding fractions lie below herd immunity, the community is at risk for additional outbreaks if all preventive measures are relaxed. This is particularly the case for the last scenario where about 15% are infected during the first outbreak.

Needless to say, there are several uncertainties in the presented predictions. One uncertainty is the time calibration which is affected by the choice *s_D_* = 21 (typical time between infection and death) and the infection fatality risk *f* = 0.3%. However, changing *s_D_* say 4 days up or down only shifts the time calibration by the same number of days, and changing *f* to 0.1% or 1% only moves the time calibration by less than a week forward or backward.

The initial doubling time *d* = 3.5, and even more so the new doubling time after prevention, play a more significant role, more so than changing *R*_0_ = 2.5 by 10% up or down. If the initial doubling time instead was set to *d* = 4 this would have slowed down the entire epidemic by about a week. The big effect of having different doubling times after preventive measures are put in place, corresponding to different magnitude of preventive measures, is illustrated by the alternative predictive curves in the figures.

### 4 Additional comments

The underlying epidemic model was the simple SIR General epidemic model. This model only allows for heterogeneity in terms of infectiousness (when and how many to infect), but not in terms of susceptibility or mixing-patterns. To include such heterogeneities is of course important and often done (e.g. [5],[3]). The purpose of the present paper is however to keep things simple enough in order to make procedures more transparent. Another feature in the model that would make it more realistic is to include a latent state before becoming infectious by instead using SEIR models [4].

When comparing effects of prevention one can either compare the reduction in final number getting infected, and/or the size of the incidence peak since this is when health burden is most problematic for the health care system. As an illustration, assuming Stockholm follows the green curve, the final number getting infected is reduced from 78% (with no preventive measures) to 47%, a reduction of 40%. The peak height is reduced from 59 000 infected on the peak day to 17 000 on peak day, a reduction of about 70%. To compare the number of infections at a given time point during the outbreak is however not a sensible comparison, since the more preventive scenarios have their infections shifted to the right (cf. [6]). As an illustration, comparing the black and green curve of Figure 2 near the peak (e.g. looking at April 1) the fraction infected in the black curve equals 59%, and in the green curve 22%, which indicates that the number of infected is reduced by about 62%. There will however be more infections to come in the preventive curve, and at the end of the outbreak the reduction is 40%.

There are several papers doing more advanced and realistic modelling/prediction (e.g. [3], [6], [10]). However, our estimation-prediction methodology is much simpler and straight-forward to implement, and we feel it is a useful complement to the more advanced methods referred to. As a consequence, it is much more transparent to see how the few model assumptions affect the results, and it is easy to vary the few parameters to see their effect on predictions. We hope the method will increase understanding about which parameter-uncertainties that have biggest impact on predictions, and which parameter-uncertainties that are less influential. Finally, we expect this simple method to give predictions being quite similar to the more complicated models, and if they don’t there is strong reasons to investigate why this is not the case.

There are of course also obvious advantages with more realistic models containing e.g. age-structure, households, work places, symptom response and different preventive measures. Spatial aspects are less important since we consider one city-urban area. More advanced models will give better fit if correct parameter estimates are used, and also more questions can be addressed, such as infection risk in different age-groups, which means of spread is most common, and the effectiveness of different preventive measures as well as when making statements about case fatalities and hospitalization. The general effect of making a more realistic model with more heterogeneities is that slightly fewer will get infected and that the peak height is slightly lowered and shifted a week or two later.

## References

1. Ferguson Laydon, Nedjati-Gilani et al. (2020). Impact of non-pharmaceutical interventions (NPIs) to reduce covid-19 mortality and healthcare demand. Imperial College covid-19 Response Team, March 16, 2020.

2. Sjödin, H., Johansson, A.F., Brännström, Å., Farooq, Z., Kriit, H.K., Wilder-Smith, A., Åström, C., Thunberg, J. and Rocklöv, J. (2020). Covid-19 healthcare demand and mortality in Sweden in response to non-pharmaceutical (NPIs) mitigation and supression scenarios. MedRxiv https://doi.org/10.1101/2020.03.20.20039594

3. Diekmann O., Heesterbeek, J.A.P. and Britton, T. (2013). Mathematical tools for understanding infectious disease dynamics. Princeton UP.

4. Swedish Public Health Agency, www.folhalsomyndigheten.se

5. Li, R., Pei, S., Chen, B., Song, Y., Zhang, T., Yang, W.\ and Shaman, J. (2020). Substantial undocumented infection facilitates the rapid dissemination of novel coronavirus (SARS-CoV2). Science. 10.1126/science.abb3221

6. Swedish Peublic Health Agency. Skattning av peakdag och antal infekterade I covid-19-utbrottet I Stockholms län februari-april 2020. Official preprint. https://www.folkhalsomyndigheten.se/publicerat-material/publikationsarkiv/s/skattning-av-peakdag-och-antal-infekterade-i-covid-19-utbrottet-i-stockholms-lan-februari-april-2020/

7. Flaxman, S., Mishra, S., Gandy, A. et al. (2020). Estimating the number of infections and the impact of non-pharmaceutical interventions on covid-19 in 11 European countries. Imperial College covid-19 Response Team, March 30, 2020.

8. Wu, J.T., Leung, K., Bushman, M., Kishore, N., Niehus, R., de Salazar, P.M., Cowling, B.J., Lipsitch, M. and Leung, G.M. (2020). Estimating clinical severity of COVID-19 from the transmission dynamics in Wuhan, China. \emph[Nature Medicine]. http://dx.doi.org/10.1038/s41591-020-0822-7

9. WHO (2020). Coronavirus disease (COVID-2019) situation reports. Situation report 30.

## References

[1] Britton, T. (2020) Basic prediction methodology for covid-19: estimation and sensitivity considerations. MedRxiv. https://doi.org/10.1101/2020.03.27.20045575

[2] Britton, T. and Scalia Tomba G. (2019). Estimation in emerging epidemics: biases and remedies. J. Roy. Soc. Interface. 16:20180670,

[3] Chinazzi, M., Davis, J.T., Ajelli, M., Gioannini, C., Litvinova, M., et al. (2020). The effect of travel restrictions on the spread of the 2019 novel coronavirus (COVID-19) outbreak. Science, DOI: 10.1126/science.aba9757.

[4] Diekmann O., Heesterbeek, J.A.P. and Britton, T. (2013). Mathematical tools for understanding infectious disease dynamics. Princeton UP.

[5] Ferguson Laydon, Nedjati-Gilani et al. (2020). Impacxt of non-pharmaceutical interventions (NPIs) to reduce covid-19 mortality and healthcare demand. Imperial College covid-19 Response Team, March 16, 2020.

[6] Flaxman, S., Mishra, S., Gandy, A. et al. (2020). Estimating the number of infections and the impact of non-pharmaceutical interventions on covid-19 in 11 European countries. Imperial College covid-19 Response Team, March 30, 2020.

[7] Swedish Public Health Agency, www.folhalsomyndigheten.se

[8] Li, Q., Guan, X., Wu, P., Wang, X., Zhou, L. et al. (2020). Early Transmission Dynamics in Wuhan, China, of Novel Coronavirus-Infected Pneumonia. N. Engl. J. Med. 382: 1199–1207.

[9] Li, R., Pei, S., Chen, B., Song, Y., Zhang, T., Yang, W. and Shaman, J. (2020). Substantial undocumented infection facilitates the rapid dissemination of novel coronavirus (SARS-CoV2). Science. 10.1126/science.abb3221

[10] Sjödin, H., Johansson, A.F., Brännström, Å., Farooq, Z., Kriit, H.K., Wilder-Smith, Å., Aström, C., Thunberg, J. and Rocklöv, J. (2020). Covid-19 healthcare demand and mortality in Sweden in response to non-pharmaceutical (NPIs) mitigation and supression scenarios. MedRxiv https://doi.org/10.1101/2020.03.20.20039594

[11] Wallinga, J. and Lipsitch, M. (2007) How generation intervals shape the relationship between growth rates and reproductive numbers. Proc. R. Soc. B 274: 599–604.

[12] WHO (2020). Coronavirus disease (COVID-2019) situation reports. Situation report 30.

[13] Wu, J.T., Leung, K., Bushman, M., Kishore, N., Niehus, R., de Salazar, P.M., Cowling, B.J., Lipsitch, M. and Leung, G.M. (2020). Estimating clinical severity of COVID-19 from the transmission dynamics in Wuhan, China. Nature Medicine. http://dx.doi.org/10.1038/s41591-020-0822-7

